# Feasibility of mixed-reality telecollaboration to enhance pre-medical student shadowing education

**DOI:** 10.1101/2024.09.04.24312893

**Authors:** Aleeza Nasir, Rida Nasir, Daisy Puca, Kevin Charles, Sandhya LoGalbo, Jayme Schwartz, Ishveer Kaur, Temesgen Tsige, Tran Tu Huynh, Lisa Iyeke, Lindsay Jordan, Mark Richman

## Abstract

**Introduction:** Medical school admissions are highly competitive, leading to high attrition rates, particularly among underserved minority (URM) students. Mentorship and clinical exposure are critical for the success of URM students, but limited access to these experiences can be due to factors such as time constraints and transportation barriers. Augmented reality (AR) has the potential to revolutionize medical education for pre-medical students by providing engaging and accessible clinical shadowing opportunities. This study aims to investigate the feasibility and impact of integrating OpticSurg’s Vision Beyond® AR platform into the educational experience of students shadowing the Emergency Department (ED). The study will focus on students’ experience using the Vision Beyond® platform and the potential strengths and weaknesses associated with the overall use of the device.

**Methods:** The study included Hofstra pre-medical undergraduate and post-baccalaureate students of good academic standing and character all cleared to shadow in the Long Island Jewish Medical Center Emergency Department (ED), aged 18 or older. This pilot study explored the use of Vision Beyond® AR glasses, for remote pre-medical education. Students who were not present in the ED accessed medical teaching materials using de-identified resources available on the internet, such as CT scans, EKGs, lab results, physical examination findings, ultrasounds, and X-rays. No patients were involved in the study. The educator used the Vision Beyond® goggles to start a session and invited the students to participate through the Vision Beyond® website. Students utilized the interactive feature to indicate areas where they desired greater detail or explanation. After each session, students completed a survey to assess their experience, including demographics, system usability, ease of use for specific topics, overall impression, and the strengths and weaknesses of the platform.

**Result:** Fifteen pre-medical and post-baccalaureate students participated in the study. Students rated the device on a 1-to-5 scale (1 = Strongly disagree, 5 = Strongly agree) for ease of use and learnability. On average, the ratings for these categories were 4 or above. The incidence of motion sickness and nausea received an average rating of 1.8 out of 5.

**Conclusion:** The pilot program using OpticSurg’s Vision Beyond® platform showed promise, with students reporting a positive experience and finding the device easy to use. However, challenges such as internet connectivity issues and limitations of the AR goggles were noted. Future research should explore how this technology can enhance diversity in the medical field.

## Introduction

Medical school is competitive and candidates are more likely to be admitted if they have: 1) an excellent GPA, 2) a high MCAT score, 3) mentorship, 4) research experience, and 5) volunteering experience. Unfortunately, this demanding path leads to high attrition rates, with nearly 85% of pre-med students dropping out of the track [1]. This attrition is particularly pronounced among underserved minority (URM) students [2]. For instance, mentorship plays a crucial role in supporting URM students, not only boosting their confidence in navigating the medical school application process but also offering access to resources that can help overcome existing barriers, equipping them with the skills and knowledge necessary to thrive [3]. Other factors influencing attrition include deficiencies in undergraduate advising, historical biases, and limited exposure to internships or opportunities. These factors significantly contribute to URM students being significantly underrepresented in the medical field. Increasing the number of URM students in medicine benefits society, as URM physicians are more likely to practice in underserved communities, often reflecting their ethnic background [4].

Hofstra University sponsors the course “Introduction to Clinical Research” at the Long Island Jewish Medical Center Emergency Department (ED) and many students continue to stay on for shadowing, volunteering, or research after the course ends. In addition, many non-Hofstra URM students choose to volunteer in LIJ’s ED. However, many students wishing to stay on or join for such experiences are unable to due to time commitments or being unable to come to the hospital because of transportation or commuting barriers (time and financial costs). Moreover, students from non-local schools (eg, Hopkins) are only able to benefit during short vacation times.

Augmented and virtual reality (AR/VR) devices are revolutionizing education by offering a more engaging and effective learning experience. This technology also facilitates group learning by allowing instructors to share AR/VR experiences with a large number of students simultaneously, thus providing students with a better understanding of the procedure and more accurate knowledge. Perhaps most important, AR/VR allows students to participate in virtual experiences that would be impossible or too expensive to recreate in a traditional learning environment, opening the door to a new learning paradigm [5].

To maintain URM students’ interest in careers as physicians, we hope to create the “Code Ed” program to facilitate the process of continued engagement using augmented reality via OpticSurg’s Vision Beyond® platform and hardware. Vision Beyond® ‘s goggles allow the educator to stream a (non-recorded) video to students/viewers on their phone, tablet, or desktop device. They can highlight items to focus on by touching their screen, which puts a target around the item on the educator’s screen.

The study aimed to: (1) Investigate whether OpticSurg’s Vision Beyond® platform and hardware can be integrated into the educational experience of ED shadow students. (2) Investigate whether OpticSurg’s Vision Beyond® platform and hardware are easy to use in clinical educational activities. (3) Investigate the impact of exposure to clinical shadowing via OpticSurg’s Vision Beyond® platform and hardware on students’ interest in medicine.

## Methods

This pilot program assessed the feasibility of Vision Beyond®, augmented-reality-enabled glasses, to facilitate remote pre-medical education. The study included Hofstra University’s pre-medical undergraduate or post-baccalaureate students of good academic standing and quality of character cleared to shadow in LIJ’s Emergency Department (ED), age 18 or older.

Students not in the ED had access to medical teaching using de-identified materials available on the internet: CT scans, EKGs, lab results, physical examination findings, ultrasounds, and rays. No patients were involved in the study. The educator started a session by wearing the Vision Beyond® goggles and inviting students to learn through the Vision Beyond® website. Students used the interactive feature to indicate areas for which they desired greater detail/explanation by highlighting the area with their finger directly on the screen. Teaching points for each session were chosen from among the following domains: CT, EKG, labs, physical examination, procedures (students performed alongside, via video), ultrasounds, and X-rays.

We evaluated the learner’s experience using a survey after each session that the students attended virtually. The survey assessed: 1) student demographic information, 2) 5-point Likert scale of: a) system usability, b) ease of use for each learning point, c) overall assessment and 4) student comment on the strengths and weaknesses of Vision Beyond®.

This study was deemed exempt by the Northwell Health Institutional Review Board (IRB #: 24-0580).

## Results

Fifteen pre-medical students participated in a study evaluating the Vision Beyond® device. Students rated the device on a 1-to-5 scale (1 = Strongly disagree, 5 = Strongly agree) for ease of use and learnability. On average, the rating for the categories was 4 or above. Overall, motion sickness and nausea received an average rating of 1.8 out of 5 (Figure 1).

**Figure 1.**
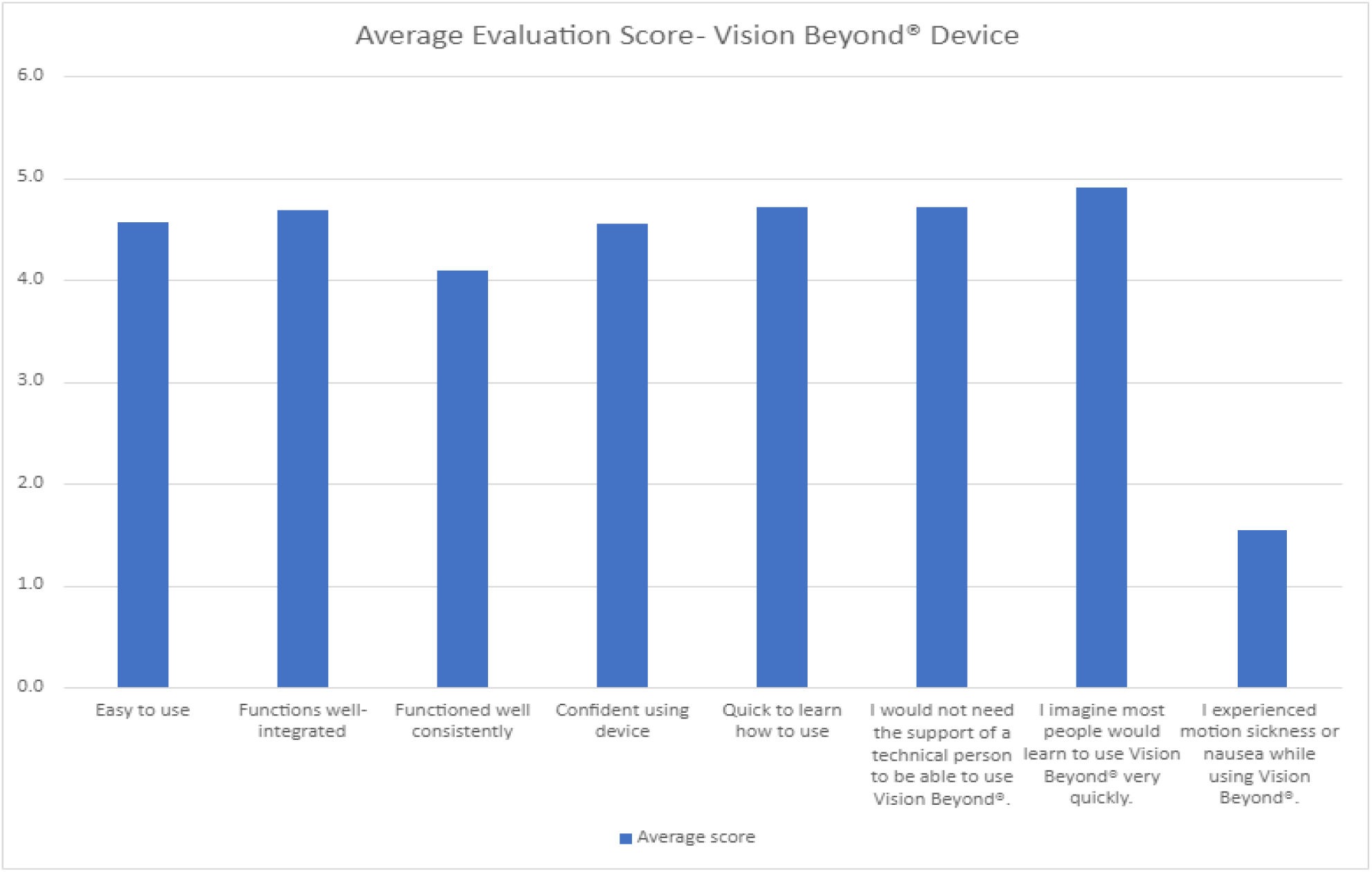
The average evaluation score for the Vision Beyond® device

Students were exposed to a variety of subject areas, including CT scans, EKGs, laboratory tests, physical examinations, procedures, ultrasounds, and X-rays. They encountered a total of 94 educational exposures **(Table 1)**.

**Table 1:** Number of students exposures by subject matter domain.

| **<u>Domain</u>** | **<u># of student exposure</u>** |
| --- | --- |
| CT | 6 |
| EKG | 3 |
| Lab | 14 |
| Physical Examination | 17 |
| Procedure | 20 |
| Ultrasound | 25 |
| X-Ray | 15 |
| **Total student-domain education exposures** | **94** |

Students provided feedback on the strengths and weaknesses of the Vision Beyond® device **(Tables 2 and 3)**.

**Table 2:** Sample Student Comments “Weaknesses”.

| <u>Weaknesses</u> |
| --- |
| Blurry screen |
| Camera moved around too much |
| Connection issues (ie., lag in audio and video from wearer to viewer) |
| Goggle camera is on top but wearer’s eyes are below them, so the camera isn’t focused on what the wearer’s eyes are focused on |
| If the wearer moves their head, the call disconnects |
| Images “blown out” (became overly white, making it difficult to see) |
| Laser pointer not working sometimes |
| Loud background noise |
| Poor image resolution |
| Procedures went by too fast to follow manually along with doing it ourselves |
| Short battery life (20-60 minutes) |
| Small visual field (wearer has to be close to the item of interest) |
| Small visual scope (ie., slides not full on the screen) |
| Sound distortion id wearer and viewers are too close |
| Target capability was occasionally lost 10 minutes into the session |

**Table 3:** Sample Student Comments “Strengths”.

| <u>Strengths</u> |
| --- |
| Allows for focus on lesson |
| Bidirectional audio communication |
| Ease-of-use |
| Easy to ask questions |
| Facilitates group learning and interaction |
| Facilitates the instruction of procedures on a large scale |
| Good for learning-by-doing procedures in real-time practice |
| Navigation and connection |
| Remote learning; synchronous |
| Strong audio and video |
| Target objects for focus |
| Time efficient and great pace of knowledge |

## Discussion

This pilot study explored the potential of mixed-reality telecollaboration via OpticSurg’s Vision Beyond® platform, to address challenges in premedical education, particularly for underserved minority (URM) students. Augmented reality (AR) has emerged as a promising tool to address these challenges by creating a more engaging learning experience, facilitating remote learning, and providing access to shadowing experiences and opportunities [6].

This pilot study successfully demonstrated the feasibility of using mixed-reality telecollaboration with OpticSurg’s Vision Beyond® platform for pre-medical education. The positive survey feedback, particularly regarding enjoyment, suggests that students found the program engaging and valuable. Furthermore, it evaluated the Vision Beyond® device and most students rated the device favorably. On a scale of 1-to-5, with 5 indicating strongly agree, most students rated the device 4 or higher for ease of use, integration of functions, functionality, confidence in using the device, and other educational domains. These results suggest that students had an overall positive experience with the platform.

Despite the program’s promising results, several challenges were identified by the students. These include: (1) occasional internet connectivity issues disrupting the live video streaming, potentially hindering the learning experience. Addressing this issue requires exploring solutions to strengthen internet stability. (2) The current version of Vision Beyond® goggles presents limitations in the user’s field of view and the range of interactive functionalities. Future versions with wider fields of view and more intuitive controls could significantly enhance the user experience and educational effectiveness.

Subsequent follow-ups should assess whether this enhanced access positively impacts the inclusion of minority students in medicine, aiming to alleviate some of the barriers they often face. Moreover, future research should go beyond the feasibility and assess the program’s long-term impact on diversity and inclusion in medicine. This could involve follow-up studies to see if the program’s enhanced access positively impacts the inclusion of minority students in the field.

## Data Availability

All data produced in the present study are available upon reasonable request to the authors.

## References

1. Zhang C, et al. The process of attrition in pre-medical studies: A large-scale analysis across 102 schools. PLoS One. 2020 Dec 28;15(12):e0243546.

2. Nguyen M, et al. Association of Sociodemographic Characteristics With US Medical Student Attrition. JAMA Intern Med. 2022 Sep 1;182(9):917–924.

3. https://www.cdc.gov/pcd/issues/2023/22_0362.htm

4. https://www.ncbi.nlm.nih.gov/pmc/articles/PMC5871929/

5. Ardiny, H., & Khanmirza, E. (2018, October). The role of AR and VR technologies in education developments: opportunities and challenges. In 2018 6th rsi international conference on robotics and mechatronics (icrom) (pp. 482–487).

6. Dhar P, Rocks T, Samarasinghe RM, Stephenson G, Smith C. Augmented reality in medical education: students’ experiences and learning outcomes. Med Educ Online. 2021 Dec;26(1):1953953. doi: 10.1080/10872981.2021.1953953. PMID: 34259122; PMCID: PMC8281102.

